# A delayed modulation of solar ultraviolet radiation on the COVID-19 transmission reflects an incubation period

**DOI:** 10.1101/2020.10.13.20183111

**Authors:** Maosheng He, Keyan Fang, Feifei Zhou, Tinghai Ou, Deliang Chen

## Abstract

Laboratory experiments have revealed the meteorological sensitivity of the coronavirus disease 2019 (COVID-19) virus. However, no consensus has been reached about how outdoor meteorological conditions modulate the virus transmission as it is also constrained by non-meteorological conditions. Here, we find that statistically, non-meteorological factors constrain the growth rate of cumulative confirmed cases least when the cases in a country arrive around 1300–3200. The least-constrained growth rate correlates with the ultraviolet flux and temperature significantly (correlation coefficients *r*=-0.55*±*0.09 and -0.40*±*0.10 at *p <* 0.01, respectively), but not with precipitation, humidity, and wind. The ultraviolet correlation exhibits a delay of about seven days, providing a meteorological measure of the incubation period. Our work reveals a seasonality of COVID-19 and a high risk of a pandemic resurgence in winter, implying a need for seasonal adaption in public policies.

**One-sentence summary:** A delayed modulation of ultraviolet radiation on the COVID-19 transmission provides independent evidence for a 7-day incubation period and implies a strong seasonality

## 1 Introduction

A way to predict the COVID-19 transmissions in winter is to quantify the responses of the virus survival and transmission to the winter weather conditions. Laboratory experiments revealed that the ultraviolet (UV) radiation and air temperature modulate the airborne survival of the virus strongly[1, 2, 3]. Meanwhile, several studies have attempted to extract relevant epidemiological evidence, e.g., [4, 5, 6, 7], through studying the correlations between the count of confirmed cases or mortality and meteorological conditions using data from priorly selected cities. Their conclusions, however, are often contradictory. Some attempts did not find the correlation, e.g., [4, 3], whereas the others reported weak or moderate correlations, e.g., [5, 6, 7]. The results are not conclusive, mainly because the transmission is constrained by non-meteorological factors which can hardly be considered in an appropriate way.

At the early stage of an uncontrolled outbreak, the infection cases grow exponentially, whereas its growth rate is relatively stable, e.g., [8]. Accordingly, here we chose the growth rate as an index of the COVID-19 transmission, which is expected to be more susceptible to meteorological conditions than the infection cases. We estimate the growth rate through a sliding window regression using the daily count of the accumulative confirmed cases from each county. Constrained by insufficient test capability at the early stage, the confirmed cases’ growth might not accurately reflect the infection growth. To deal with this problem, we calculate the correlation between the regressed growth rate and meteorological conditions across all countries as a function of the confirmed case count. The range of the accumulative confirmed cases, in which the correlation maximizes, is identified as the least-constrained stage. The growth rate at this stage is defined as the least-constrained growth rage, and its response to meteorological conditions is investigated statistically. As illustrated below, the least-constrained growth rate minimizes the contamination from non-meteorological factors, e.g., test capability and artificial controls, and allows revealing meteorological modulations in detail.

## 2 Data analyses and results

This study uses daily cumulative confirmed COVID-19 cases at a country level and daily meteorological variables until 1 September of 2020. The confirmed cases are from COVID-19 Data Repository by the Center for Systems Science and Engineering (CSSE) at Johns Hopkins University, whereas the meteorological variables are extracted from the ERA5 reanalysis dataset from the European Centre for Medium-Range Weather Forecasts (ECMWF) (C3S, 2017). The meteorological variables analyzed herein include the air temperature at the height of 2 m above the surface (land, sea or inland waters), precipitation, relative humidity, wind speed at the height of 10 m, downward UV radiation flux at the surface (UV, in the range 250-440 nm), and diurnal temperature range. The daily mean meteorological data were averaged for each country to compare with the country-level COVID-19 data. Below, we compare the growth of confirmed cases with its theoretical expectation, in an example country in Section 2.1 and statistically in Section 2.2, determine the statistical stage modulated most sensitively by weather in Section 2.3, investigate the geographic distribution of the confirmed case growth and its correlation with meteorological factors in Sections 2.4 and 2.5, and diagnose the incubation period in Section 2.6

### 2.1 Evolution of the outbreak in an example county: stages of the confirmed case growth

Fig. 1a displays the cumulative confirmed case number *y* as a function of time *t* in for Bulgaria as an example. Through a least-square regression, we fit *y* to an exponential model *y* = *ae*^*b*(*t−τ*)^ in a 28-day-wide sliding window. Here, *τ* = 86, 88, …, 232 day denotes the center of the sliding window, *a* measures the confirmed cases at *τ*, and the exponent factor *b* measures the growth rate. The regressed growth rate *b* is shown in Fig. 1b as a function of time *τ*. (Besides *b*, the regression yields also *a* and *r*^2^, a measure of the regression goodness equaling to the square of the correlation coefficient between *y*(*t*) and its regression values as used in the following sections.) The regressed growth rate *b* can be divided into three stages, labeled as I, II, and III and indicated by the blue arrows in Fig. 1. Stages I and III are characterized by decreasing growth rates, whereas Stage II is associated with a relatively stable growth rate.

**Figure 1:**
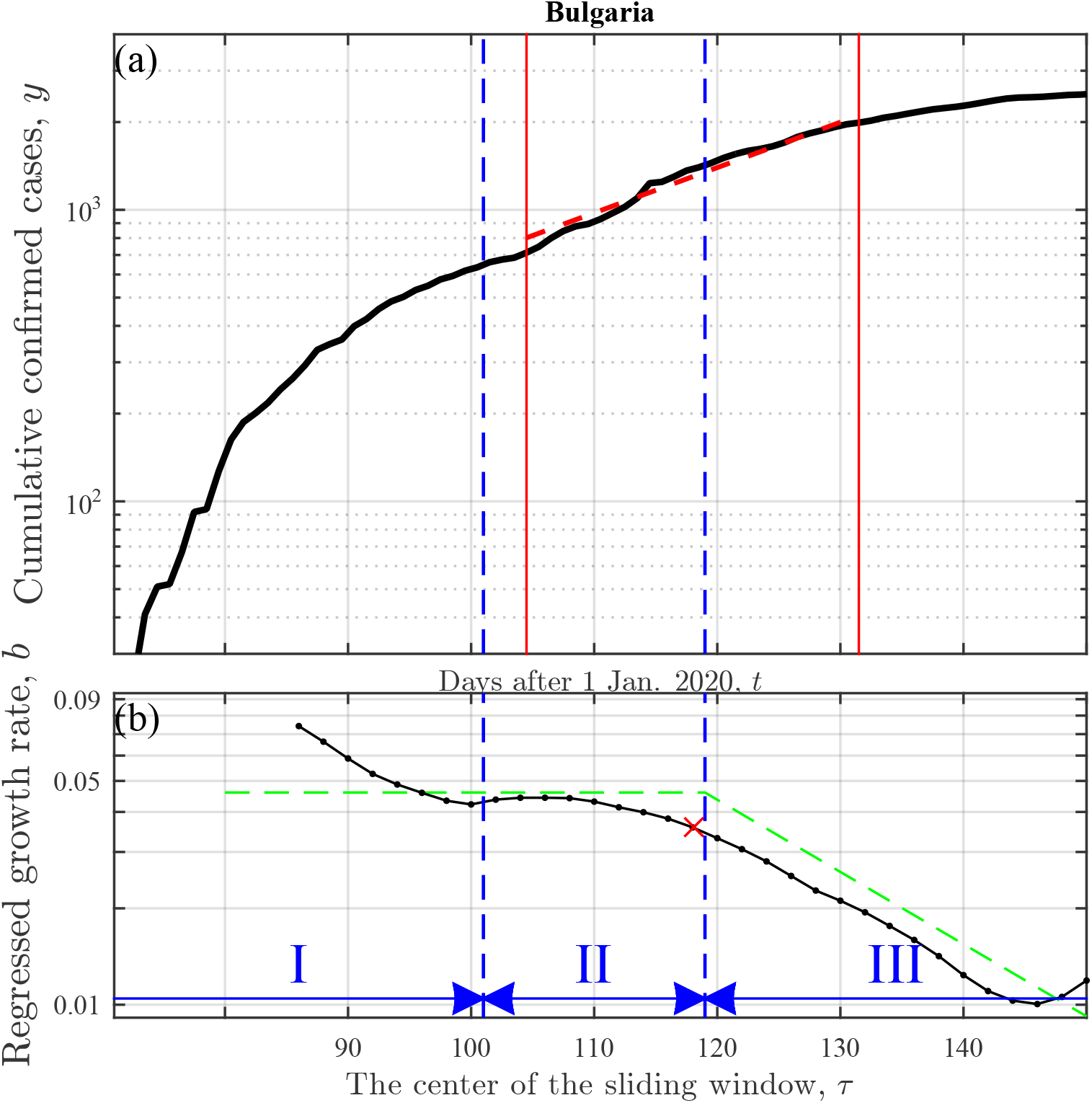
(a) Cumulative confirmed cases *y* as a function of time *t* and (b) its growth rate *b* against the regression time parameter *τ*, for Bulgaria as an example. The growth rate *b* (black line in (b)) is regressed according to the model *y* = *ae*^*b*(*t−τ*)^ in a 28-day-wide sliding window centering at *τ* =86, 88, …, 152, where *a* represents the model initial confirmed cases (namely, the regressed *y* value at *τ*). The growth of confirmed cases could be divided into three stages: Stages I, II and III, sketched by blue symbols in (b). In Stage II, the growth is stable (0.035*< b <*0.045), whereas in the other two, *b* decreases by more than 0.02. In Stages II and III but not in Stage I, the real growth is largely consistent with the ideal evolution that is sketched as the green line in (b) according to, e.g. [11, 12]. We attribute the discrepancy in Stage I to an insufficient test capability at the early stage. The red symbols denote the least-constrained growth rate and its sampling window (see Sections 2.1–2.3 for details).

Theoretically, outbreaks of infectious diseases, e.g., [8, 9, 10], are characterized by two phases: the uncontrolled first phase showing stable exponential growth followed by a second phase with a decreasing growth rate usually after effective artificial controllers [11, 12]. The two-phase theoretical growth is sketched as the green dashed line in Fig. 1b, which are largely parallel to the growth of the confirmed case (black line) in Stages II and III, and can explain largely these stages. Stage II reflects the uncontrolled transmission of COVID-19, whereas Stage III reflects the decline phase where the decreasing growth rate could be explained as responses to artificial interventions or controllers, e.g., travel restrictions and changes in human behaviors, e.g., [13, 14]. However, in Stage I in Fig. 1b, the black line is not parallel to the green. In the beginning of the outbreak, infections cannot be confirmed timely and can accumulate until sufficient tests. The decreasing growth rate of the confirmed case, biased from the theoretical growth, might reflect more the delayed sufficient test capability than the infection growth.

Compared with Stages I and III, Stage II is characterized by a relatively stable growth rate reflecting uncontrolled transmissions confirmed timely and sufficiently. Therefore, we assume that in Stage II the growth is modulated most sensitively by the weather and will use Stage II for exploring the impacts of weather. In practice, instead of delimiting Stage II for each country individually, we explore Stage II statistically for all countries in the following subsection.

### 2.2 Statistical evolution of the outbreak

The previous section defined a daily growth rate of the cumulative confirmed COVID-19 cases in an example country through regression and used the growth rate to describe the evolution of the outbreak. We further implement the regression for every country, yielding *a, b*, and *r*^2^ as functions of *τ*. Fig. 2a and 2b display *b* against *a* and *r*^2^ from the 50% most developed countries, respectively. These countries are characterized by the gross domestic product per capita 2019 above the median of all countries, 6200 United States dollars, and therefore are presumably least subject more to the socioeconomic factors [15, 16] as discussed further in Section 2.6. Here, we use *a* rather than the date *t* to coordinate the growth rate since the outbreaks do not occur at the same date in different countries. In Fig. 2b, *b* exhibits a dependence on *r*^2^ at *r*^2^ *<*0.9. Accordingly, we exclude *b* values associated with *r*^2^ *<*0.9 from the following analyses.

**Figure 2:**
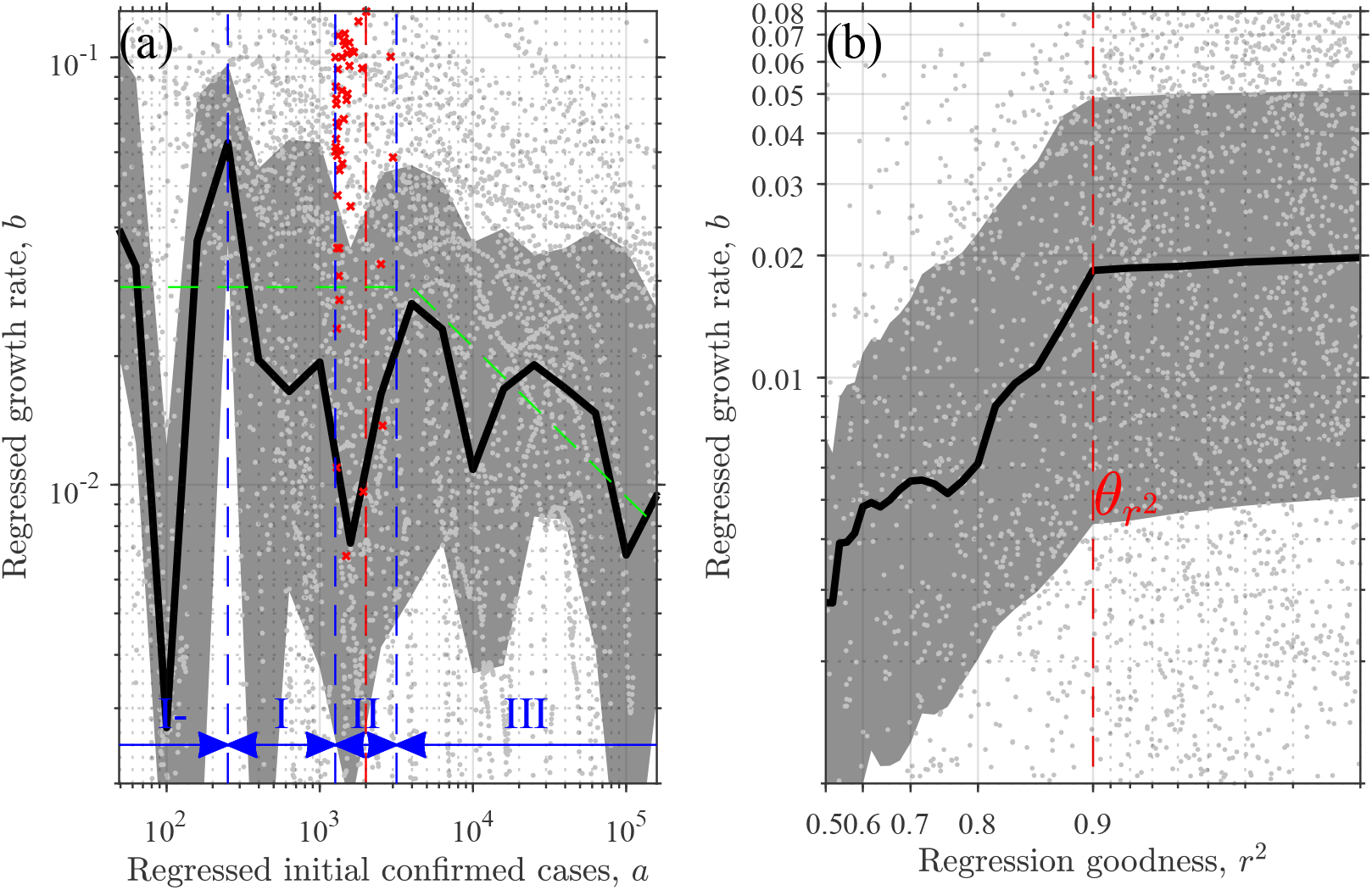
The growth rate *b* of the confirmed cases against (a) the regressed initial confirmed cases *a* and (b) the goodness *r*^2^ of the exponential regression as instanced in Fig. 1. In each panel, one dot corresponds to one sliding step of the regression; and the bold black solid line and gray shadow illustrate the median and the interquartile range, respectively. In (a), the green line sketches the ideal evolution according to, e.g. [12]; the blue symbols sketch different Stages of the evolution of the outbreak among which Stages I, II and III are the statistical estimations of Stages I, II and III instanced in Fig. 1. Stage II is centering at the red dashed line *a*_*c*_ =2000, determined in Section 2.3. In (a), the red crosses denote the least-constrained growth rate *b*_*m*_, namely, the maximum *b* in Stage II for each country. In (b), the red line displays the threshold value 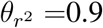 in (b) used in selecting *b*_*m*_. See Section 2.2 for the details.

In Fig. 2a, the growth curve *b*(*a*) does not clearly exhibit the three-stage shape of *b*(*τ*) in Fig. 1b but looks more complicated. The curve minimizes at *a*=10^2^, 10^3^–2*×*10^3^, 10^4^ and 10^5^. These minima might be signatures of data manipulation, e.g., [17], that some governments might try to “control” the infection cases to cross the orders of magnitude by manipulating the data. Neglecting these minima, we divide largely the growth curve *b*(*a*) into four stages. The final stage is characterized by a decreasing rate *b*(*a*), corresponding to Stage III of the *b*(*τ*) growth in Fig. 1b, and is labeled here also as Stage III. The other stages of *b*(*a*) are labeled as I-, I, and II, respectively. Stage I- is associated with the small number of infection at an the very beginning, and the growth might be susceptible to uncertainties and therefore is excluded from the following analyses. Stage I is characterized largely by a decreasing growth rate *b*(*a*), corresponding to Stage I of *b*(*τ*) in Fig. 1b. We assume that the *b*(*a*) in-between Stages I and III correspond to Stage II in Fig. 1b where the growth is relatively stable and reflects timely and sufficiently confirmed uncontrolled transmissions. Assuming that in Stage II the growth is modulated most sensitively by weather, we determine the statistical range of Stage II in the following subsection.

### 2.3 The statistical stage modulated most sensitively by weather

The statistical Stage II can be denoted as *a*_*c*_ *· S*^*−*1^ *< a < a*_*c*_ *· S* where *S* represents the half-width of the stage and *a*_*c*_ represents the stage center. We temporally assign *S*=10^0.2^ and *a*_*c*_= 10^2^, search the maximum *b*(*a*) in the range *a*_*c*_ *· S*^*−*1^ *< a < a*_*c*_ *· S* for each country, and calculate its correlation coefficient *r*_*UV*_ with the UV flux (as detailed in the following subsections) across all countries. Similarly, we search the maximum correlation coefficient *r*_*UV*_ in *a*_*c*_ *·S*^*−*1^ *< a < a*_*c*_ *·S* for *a*_*c*_= 10^2.1^,10^2.2^,…, and 10^4.0^, at *S*=10^0.2^. The absolute value |*r*_*UV*_ | as a function of *a*_*c*_ is displayed as the solid blue line in Fig. 3, which maximizes between *a*_*c*_=2000–4000. The maximum implies that at this stage of *a* the growth is modulated statistically strongest by the meteorological conditions.

**Figure 3:**
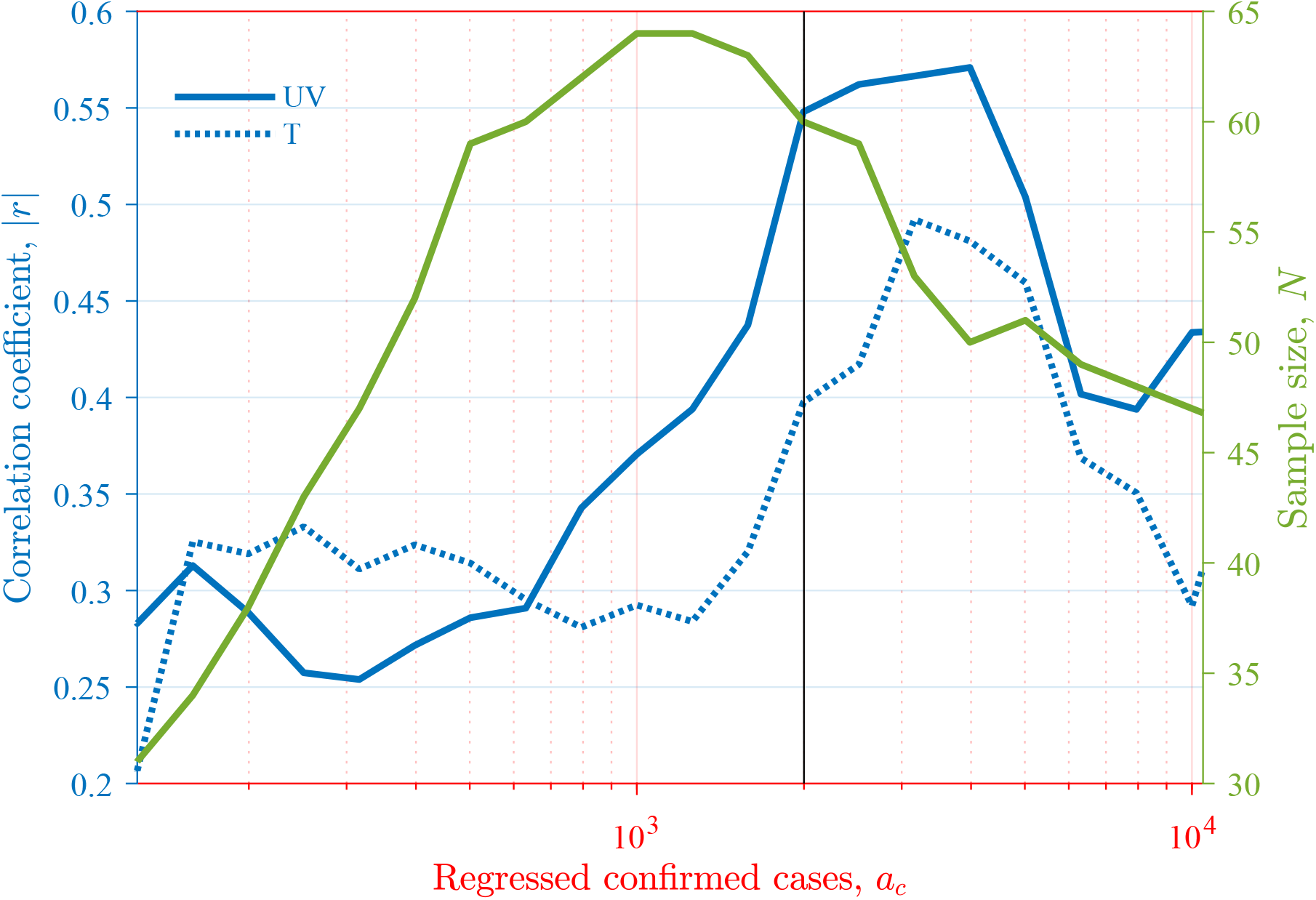
The absolute correlation coefficients |*r*| (between *b*_*m*_ and two meteorological factors) and valid sampling size *N* as functions of *a*_*c*_. The black line represents the *a*_*c*_ used for calculating the *b*_*m*_ in the current work. See Section 2.3 for the details.

Therefore, we take 2000 *×* 10^*−*0.2^ *< a <* 2000 *×* 10^0.2^ (approximately 1300*< a <*3200) as the statistical Stage II. In this *a* range, the maximum growth rate *b*(*a*) in each country is identified, referred to as the least-constrained growth rate *b*_*m*_. The red crosses in Fig. 2a display the *b*_*m*_ of all countries, associated with a median *a*=1424.

Note that the conclusions of the current work are not subject to our chooses *S*=10^0.2^ and *a*_*c*_ = 2000, learn from the same Figures as presented in the current work but for *S*=10^0.1^ and 10^0.3^, and for *a*_*c*_=1500 and 2500 (not shown).

### 2.4 Geographical distribution of the daily growth rate in percentage

According to our regression model, the least-constrained growth rate *b*_*m*_ is an exponent and 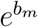 measures the ratio of the regressed number of confirmed cases of one day over that of the previous day. Therefore, 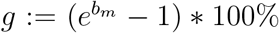 is the daily growth rate by percentage. When *b*_*m*_ *≈*0, *b*_*m*_ is already a first-order approximation of *g* due to 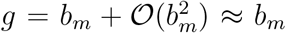, since *e*^*b*^ can be expanded into Taylor polynomial 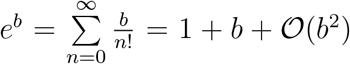. Here, *𝒪*(*b*^2^) denotes a variable with absolute value at most some constant times |*b*^2^| when *b* is close enough to 0. The following analyses are based on *g*. The geographical distribution of *g* is shown in Fig. 4. The growth rate exhibits obvious spatial clusters, e.g., the distinct difference between east and west Europe.

**Figure 4:**
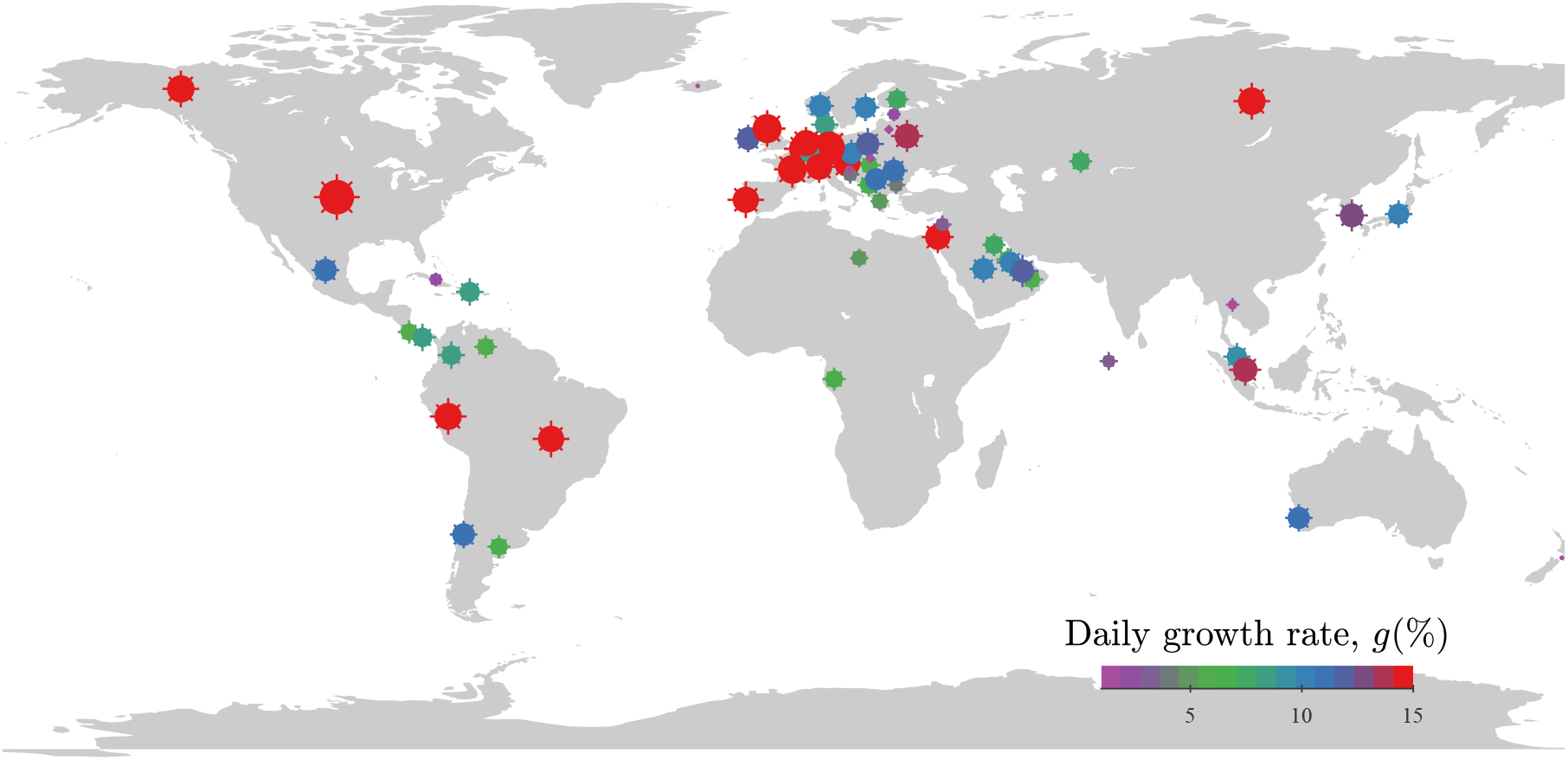
Global distribution of the daily growth rate *g* of COVID-19 confirmed cases. Each point represents one country/region. Both the color and the size of the symbols represent the growth rate. The growth rate is estimated through a sliding window regression detailed in Section 2.1 and an optimization in Section 2.3. Here, only 50% of most developed countries are included (according to the gross domestic product per capita 2019).

### 2.5 Correlations of the daily growth rate with the meteorological factors

Fig. 5 presents the correlation analyses between the least-constrained growth rate *g* and six meteorological factors, i.e., (a) the ultraviolet (UV) flux in the range 250-440nm, (b) the air temperature at 2m above the surface, (c) the diurnal temperature range, (d) the relative humidity, (e) the wind speed at the height of 10m, and (f) the precipitation. The meteorological factors are sampled and averaged in a 28-day-wide window centering at 7 days before the center of the growth rate’s sampling window. The 7-day displacement is used to deal with the COVID-19 incubation period, e.g., [18]. We determine the length of the incubation period in the following subsection.

**Figure 5:**
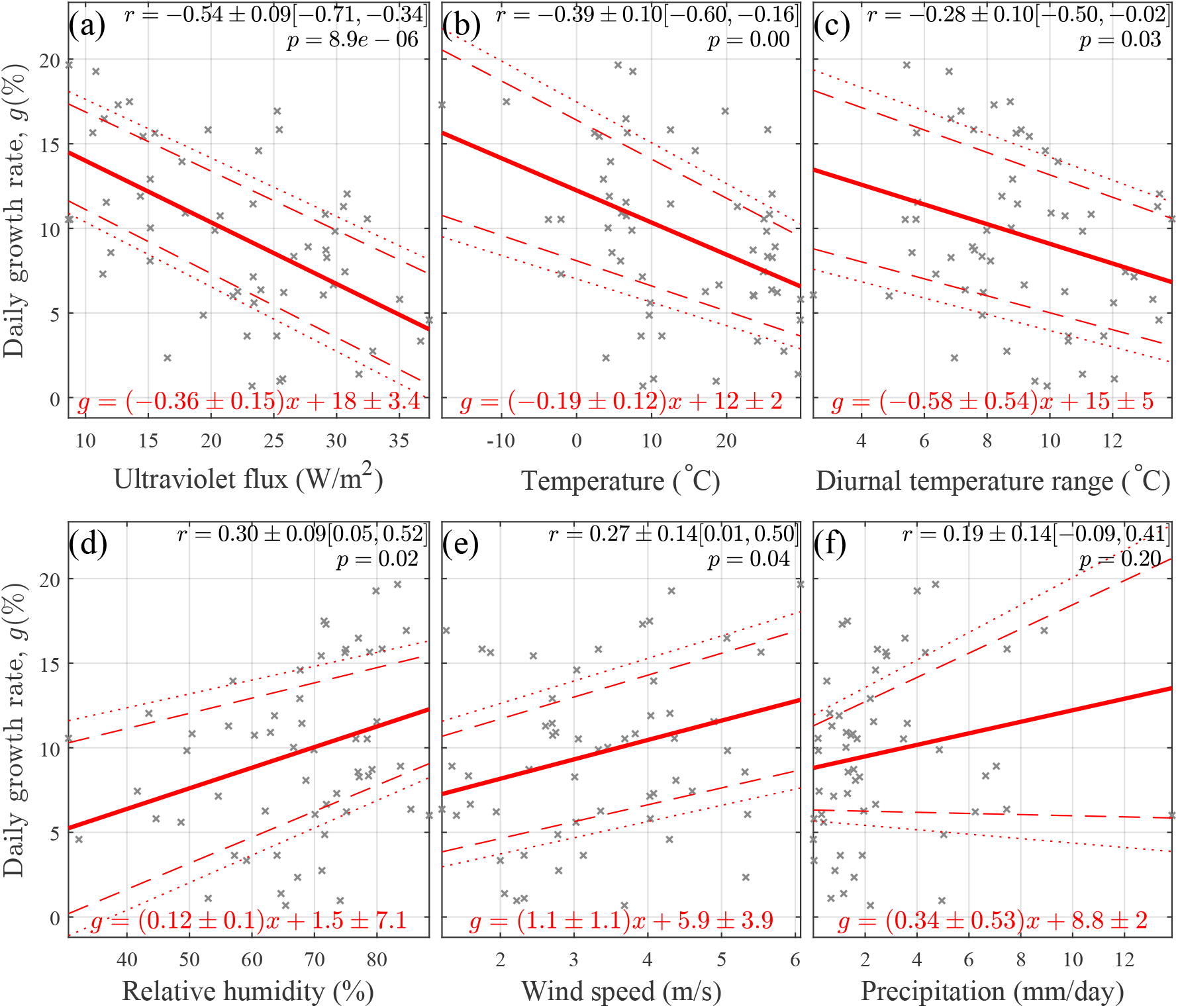
Correlation between the daily growth rate *g* and six meteorological variables: (a) the ultraviolet (UV) flux in the range 250-440nm, (b) the air temperature at 2m above the surface, (c) the diurnal temperature range, (d) the relative humidity, (e) the wind speed at the height of 10m, and (f) the precipitation. In each panel, one cross represents one country, corresponding to one red cross displayed in Fig. 2a; the solid red line presents a robust regression to a linear model *g* = *β*_1_ *\* x* + *β*_0_ through the least absolute deviations method, and the dashed and dotted lines display the significance level *α* = 0.05 and 0.01, respectively. Here, *x* denotes one of the above six variables, and *β*_1_ and *β*_0_ denote the parameters to be determined. The regression results are displayed in red on the bottom of each panel, while the Pearson correlation coefficient *r* is printed on the upright conner, in the format of *r* ± Δ*r*[*r*_*l*_, *r*_*u*_]. Here, *r* and Δ*r* are the mean coefficient and its standard deviation estimated through a bootstrapping method, and *r*_*l*_ and *r*_*u*_ are the lower and upper bounds for a 95% confidence interval. Also displayed on the top is the *p*-value for testing the hypothesis of no correlation.

The growth rate exhibits a significant correlation with the UV flux and the air temperature (*r*=-0.55*±*0.09 and -0.40*±*0.10 at *p <<*0.01, in Fig. 5a and 5b, respectively), but not with the other meteorological conditions, namely, wind speed, relative humidity, diurnal temperature range, and precipitation (*p >*=0.05, Fig. 5c–5f). According to the regressions, an increase in UV flux by 1 W/m^2^ is associated with a decrease in the growth rate by 0.36*±*0.15% per day, and an increase of the temperature by 1°C is associated with a decrease in the growth rate by 0.19*±*0.12% per day.

### 2.6 A cross-correlation analysis suggests an incubation period

The previous subsection uses a time displacement *δt* = 7day between the sampling window of the growth rate and that of the meteorological factors to deal with the incubation period. To determine the incubation period, we calculate the absolute correlation coefficient of the growth rate with the UV flux |*r*_*UV*_ |, as a function of the displacement *δt*, displayed in Fig. 6.

**Figure 6:**
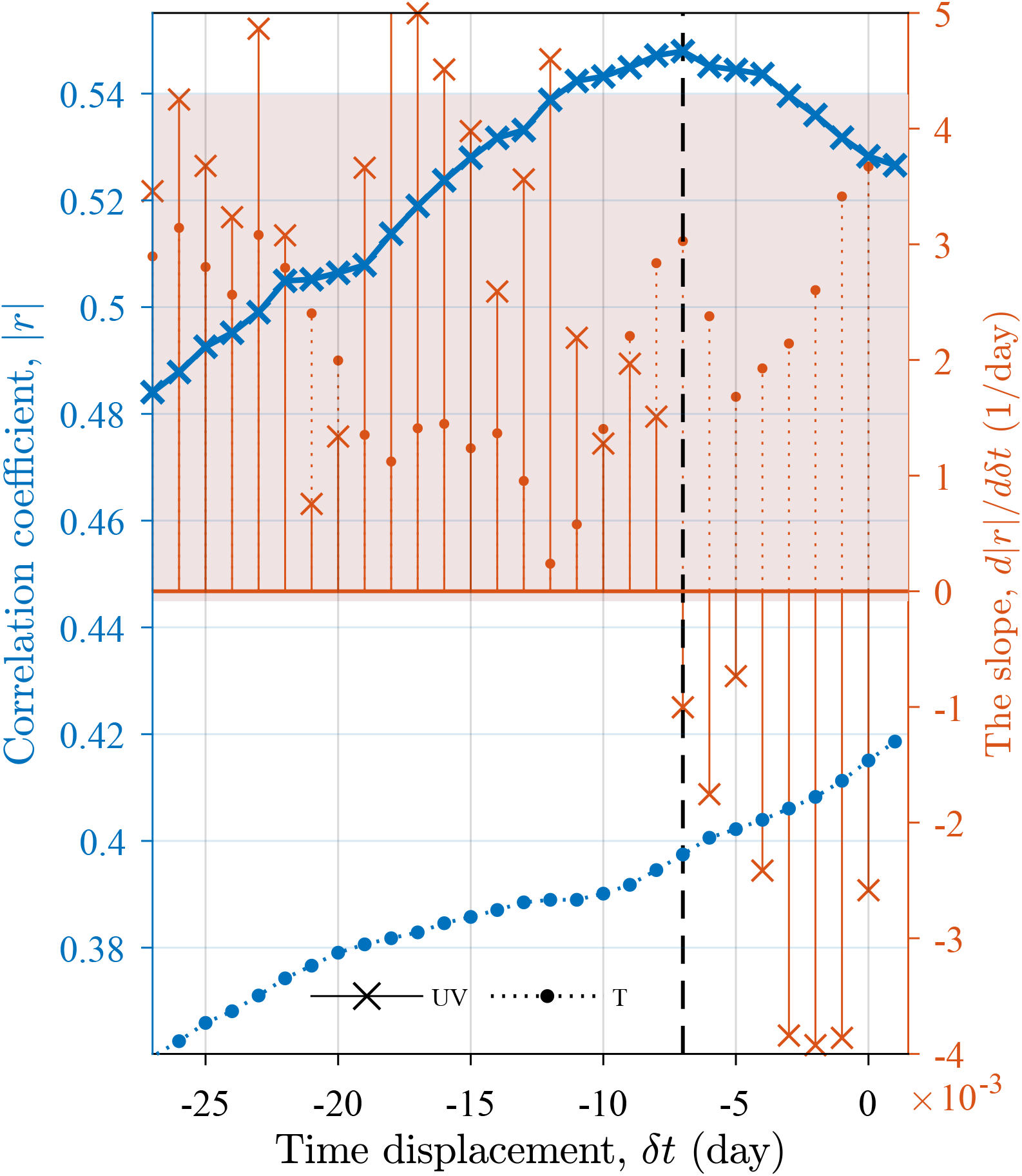
The absolute correlation coefficients |*r*| between the growth rate *g* and two meteorological factors, the UV flux and temperature *T*, as functions of the time displacement *δt*, and their slopes calculated using the centered differencing method. The shadow illustrates one standard deviation of the slopes below and above their average. See Section 2.6 for the details.

In Fig. 6, |*r*_*UV*_ | maximizes at a displacement of seven days, indicating a 7-day delayed response of the growth to UV flux. A clinical study [19] suggested a mean of an incubation period of 6.4 days, while a cross-sectional and a forward follow-up analysis [16] reported a median incubation of 7.76 days. These measures of the incubation period are comparable to the 7-day delay diagnosed herein. Our results provide evidence of the incubation period from analyzing confirmed cases instead of controlled experiments from clinical studies.

All results presented above are based on 50% of the most developed countries. For comparison, we conducted the same investigations with data from all countries (not shown), and found that the incubation period signature weakened. The weakening could be attributed to the socioeconomic factors and potentially less timely tests in less developed countries, e.g., [15, 16]. The delayed tests could reshape the cross-correlation analysis and bias the quantification of the meteorological modulation.

## 3 Discussions and conclusions

Above we illustrate that the least-constrained growth rate exhibits obvious spatial clusters, significant correlations with the meteorological conditions (UV flux and temperature). The UV correlation exhibits a delay of about seven days, which is explained as a signature of the incubation period. While spatial clusters and the correlation might be attributed to socioeconomic factors’ spatial difference [15, 16], the delay cannot. The temporal variations of socioeconomic factors are at scales much longer than the 7-day delay, which, therefore, can neither modulate the COVID-19 nor respond to the UV flux at seven days. On the other way around, the presence of the incubation period signature indicates the robustness of the UV modulation on transmission.

There are at least three factors through which meteorological conditions can modulate the transmission. The first is human behaviors. When the temperature is low, humans typically spend more time indoors, with reduced social distancing and less ventilation than outdoors. As an example, schools are places of enhanced influenza transmission [20] for intense indoor activities. The second factor is the immune system of susceptible hosts. Solar radiation drives changes in the human immune system by modulating melatonin [21] and/or vitamin D [22, 23, 24].

The last but might be the most important factor is the virus’s survival, namely the UV’s virucidal effect. Evidence has revealed that aerosols are a medium of transmission of COVID-19, as the virus remains active on the surfaces for several hours to days [11]. Intense solar radiation may inactivate the virus on the surface through the physical properties (i.e., shape, size) and the virus’s genetic material [25, 2, 26]. Simulation results revealed that 90% of the virus could be inactive under summer daytime for 6 minutes, whereas the virus becomes inactive for 125 minutes under night conditions [1]. In addition, high temperature shortens the virus survival time [27, 28, 3]. On the opposite, low temperature favors prolonging survival on infected surfaces and aerosols, which promotes the diffusion of the infection. The modulation of relative humidity, on the other hand, is negligible, as supported by laboratory experiments[1], which is different from the sensitive modulation on the influenza virus survival [29] and transmission [30].

The 7-day-delayed response to the UV flux (Fig. 6) reflects the incubation period, whereas the temperature response does exhibit a delay. A potential explanation is that temperature variation is characterized by a temporal scale longer than the incubation period, and therefore cannot resolve the incubation period. Another potential explanation is that the temperature might not be an independent drive of the transmission but a response to solar radiation. The temperature correlates significantly with the UV flux (*r*=0.86*±*0.03 at *p*=1.3*×*10^*−*17^). We carried out a canonical-correlation analysis, e.g., [31], between the growth rate and the UV flux and temperature, resulting in a canonical correlation coefficient *c*_*UV,T*_ =-0.58*±*0.08. The canonical correlation coefficient is close to the UV correlation coefficient *r*_*UV*_ =-0.55*±*0.09 (Fig. 5a), which means that using both the UV and temperature as predictors can not explain more variance than using the UV alone.

The UV impact can drive a seasonality of COVID-19 transmission and explain the following geographic dependence of COVID-19. (1) The mortality exhibits a latitudinal dependence [24]. (2) The late outbreak in Africa and arid central Asia is attributable to intense UV flux due to the low cloud fraction prior. (3) The onset of the Asian summer monsoon, increases clouds in early May [32] and yields low UV flux, which may account for the late outbreak in India and many southeastern Asia countries until early May. (4) The decrease in UV and temperature during the coming austral winter can contribute to the sharp increase in South America. For example, both the confirmed and dead cases in Brazil ranked second in the world since 13 June, and the transmission enhanced at high latitudes, such as North America and Europe November 2020.

The current study provides evidence to support the hypothesis that the UV radiation and air temperature drive the COVID-19 transmission [24]. Our results also imply a seasonality of COVID-19 and provide a meteorological measure of the incubation period. The virus transmits more readily during winter and during the global monsoon season, which impacts about 70% of the global population [33]. Accordingly, we predict a high possibility of a resurgence in the boreal winter and suggest adapting the public policy according to the seasonal variability.

## Data Availability

We used the COVID-19 data of accumulated confirmed cases until 20 July of 2020 at a country level from COVID-19 Data Repository by the Center for Systems Science and Engineering (CSSE, https://github.com/CSSEGISandData/COVID-19) at Johns Hopkins University.The daily meteorological variables are extracted from the ERA5 reanalysis dataset from the European Centre for Medium-Range Weather Forecasts (ECMWF) (C3S, 2017, https://apps.ecmwf.int/datasets/data/interim-full-daily/levtype=sfc/). The meteorological variables analyzed herein include the air temperature at 2m above the surface (land, sea or inland waters), precipitation, relative humidity, wind speed at the height of 10m,
downward UV radiation flux at the surface (UV, in the range 250-440 nm),
and diurnal temperature range. The daily mean meteorological data were averaged for each country to compare with the country-level COVID-19 data.

https://apps.ecmwf.int/datasets/data/interim-full-daily/levtype=sfc/

## Acknowledgments

This study was funded by the National Science Foundation of China (41888101, 41822101 and 41971022), Strategic Priority Research Program of the Chinese Academy of Sciences (XDB26020000), the State Administration of Foreign Experts Affairs of China (GS20190157002), fellowship for the National Youth Talent Support Program of China (Ten Thousand People Plan). Support from the Swedish Formas (Future Research Leaders) project is also acknowledged. We used the COVID-19 data of cumulative confirmed cases until 1 September of 2020 at a country level from COVID-19 Data Repository by the Center for Systems Science and Engineering (CSSE, https://github.com/CSSEGISandData/COVID-19) at Johns Hopkins University. The daily meteorological variables are extracted from the ERA5 reanalysis at C3S Climate Data Store, see https://doi.org/10.24381/cds.adbb2d47 (C3S, 2017). The gross domestic product per capita 2019 data are download from the World Bank (https://data.worldbank.org/).

## References

[1] M. Schuit, S. Ratnesar-Shumate, J. Yolitz, G. Williams, W. Weaver, B. Green, D. Miller, M. Krause, K. Beck, S. Wood, B. Holland, J. Bohannon, D. Freeburger, I. Hooper, J. Biryukov, L. A. Altamura, V. Wahl, M. Hevey, P. Dabisch, Airborne SARS-CoV-2 Is Rapidly Inactivated by Simulated Sunlight. J. Infect. Dis. 222, 564–571 (2020).

[2] S. Ratnesar-Shumate, G. Williams, B. Green, M. Krause, B. Holland, S. Wood, J. Bohannon, J. Boydston, D. Freeburger, I. Hooper, K. Beck, J. Yeager, L. A. Altamura, J. Biryukov, J. Yolitz, M. Schuit, V. Wahl, M. Hevey, P. Dabisch, Simulated Sunlight Rapidly Inactivates SARS-CoV-2 on Surfaces. J. Infect. Dis. 222, 214–222 (2020).

[3] M. Ujiie, S. Tsuzuki, N. Ohmagari, Effect of temperature on the infectivity of COVID-19. Int. J. Infect. Dis. 95, 301–303 (2020).

[4] Y. Yao, J. Pan, Z. Liu, X. Meng, W. Wang, H. Kan, W. Wang, No association of COVID-19 transmission with temperature or UV radiation in Chinese cities. Eur. Respir. J. 55, 2000517 (2020).

[5] H. Qi, S. Xiao, R. Shi, M. P. Ward, Y. Chen, W. Tu, Q. Su, W. Wang, X. Wang, Z. Zhang, Covid-19 transmission in mainland china is associated with temperature and humidity: A time-series analysis. Science of The Total Environment 728, 138778 (2020).

[6] M. M. Sajadi, P. Habibzadeh, A. Vintzileos, S. Shokouhi, F. Miralles-Wilhelm, A. Amoroso, Temperature, Humidity, and Latitude Analysis to Estimate Potential Spread and Seasonality of Coronavirus Disease 2019 (COVID-19). JAMA Netw. open 3, e2011834–e2011834 (2020).

[7] M. M. Iqbal, I. Abid, S. Hussain, N. Shahzad, M. S. Waqas, M. J. Iqbal, The effects of regional climatic condition on the spread of COVID-19 at global scale. Sci. Total Environ. 739, 140101 (2020).

[8] B. F. Maier, D. Brockmann, Effective containment explains subexponential growth in recent confirmed COVID-19 cases in China. Science (80-.). 368, 742–746 (2020).

[9] S. d. Picoli Junior, J. J. V. Teixeira, H. V. Ribeiro, L. C. Malacarne, R. P. B. d. Santos, R. d. S. Mendes, Spreading patterns of the influenza a (h1n1) pandemic. PLOS ONE 6, 1–4 (2011).

[10] A. G. Hunt, Exponential growth in ebola outbreak since may 14, 2014. Complexity 20, 8–11 (2014).

[11] Y. Liu, A. A. Gayle, A. Wilder-Smith, J. Rocklöv, The reproductive number of COVID-19 is higher compared to SARS coronavirus. J. Travel Med. 27 (2020).

[12] R. M. Anderson, B. Anderson, R. M. May, Infectious diseases of humans: dynamics and control (Oxford university press, 1992).

[13] S. Lai, N. W. Ruktanonchai, L. Zhou, O. Prosper, W. Luo, J. R. Floyd, A. Wesolowski, M. Santillana, C. Zhang, X. Du, H. Yu, A. J. Tatem, Effect of non-pharmaceutical interventions to contain COVID-19 in China. Nature (2020).

[14] H. Tian, Y. Liu, Y. Li, C. H. Wu, B. Chen, M. U. Kraemer, B. Li, J. Cai, B. Xu, Q. Yang, B. Wang, P. Yang, Y. Cui, Y. Song, P. Zheng, Q. Wang, O. N. Bjornstad, R. Yang, B. T. Grenfell, O. G. Pybus, C. Dye, An investigation of transmission control measures during the first 50 days of the COVID-19 epidemic in China. Science (80-.). 368, 638–642 (2020).

[15] S. Khalatbari-Soltani, R. C. Cumming, C. Delpierre, M. Kelly-Irving, Importance of collecting data on socioeconomic determinants from the early stage of the COVID-19 outbreak onwards. J. Epidemiol. Community Health 74, 620–623 (2020).

[16] A. Guha, J. Bonsu, A. Dey, D. Addison, Community and Socioeconomic Factors Associated with COVID-19 in the United States: Zip code level cross sectional analysis. medRxiv Prepr. Serv. Heal. Sci. p. 2020.04.19.20071944 (2020).

[17] M. Kapoor, A. Malani, S. Ravi, A. Agrawal, Authoritarian governments appear to manipulate covid data (2020).

[18] S. A. Lauer, K. H. Grantz, Q. Bi, F. K. Jones, Q. Zheng, H. R. Meredith, A. S. Azman, N. G. Reich, J. Lessler, The incubation period of coronavirus disease 2019 (CoVID-19) from publicly reported confirmed cases: Estimation and application. Ann. Intern. Med. 172, 577–582 (2020).

[19] J. Qin, C. You, Q. Lin, T. Hu, S. Yu, X.-H. Zhou, Estimation of incubation period distribution of covid-19 using disease onset forward time: A novel cross-sectional and forward follow-up study. Science Advances 6 (2020).

[20] S. Cauchemez, A. J. Valleron, P. Y. Boëlle, A. Flahault, N. M. Ferguson, Estimating the impact of school closure on influenza transmission from Sentinel data. Nature 452, 750–754 (2008).

[21] S. F. Dowell, Seasonal variation in host susceptibility and cycles of certain infectious diseases. Emerg. Infect. Dis. 7, 369–374 (2001).

[22] Abhimanyu, A. K. Coussens, The role of uv radiation and vitamin d in the seasonality and outcomes of infectious disease. Photochem. Photobiol. Sci. 16, 314–338 (2017).

[23] A. R. Martineau, D. A. Jolliffe, R. L. Hooper, L. Greenberg, J. F. Aloia, P. Bergman, G. Dubnov-Raz, S. Esposito, D. Ganmaa, A. A. Ginde, E. C. Goodall, C. C. Grant, C. J. Griffiths, W. Janssens, I. Laaksi, S. Manaseki-Holland, D. Mauger, D. R. Murdoch, R. Neale, J. R. Rees, S. Simpson, I. Stelmach, G. T. Kumar, M. Urashima, C. A. Camargo, Vitamin d supplementation to prevent acute respiratory tract infections: systematic review and meta-analysis of individual participant data. BMJ 356 (2017).

[24] P. B. Whittemore, COVID-19 Fatalities, Latitude, Sunlight, and Vitamin D. Am. J. Infect. Control (2020).

[25] J. L. Sagripanti, C. D. Lytle, Inactivation of influenza virus by solar radiation. Photochem. Photobiol. 83, 1278–1282 (2007).

[26] D. Sutton, E. W. Aldous, C. J. Warren, C. M. Fuller, D. J. Alexander, I. H. Brown, Inactivation of the infectivity of two highly pathogenic avian influenza viruses and a virulent Newcastle disease virus by ultraviolet radiation. Avian Pathol. 42, 566–568 (2013).

[27] J. M. Abduljalil, B. M. Abduljalil, Epidemiology, genome, and clinical features of the pandemic SARS-CoV-2: a recent view. New Microbes New Infect. 35 (2020).

[28] S. S. Gunthe, B. Swain, S. S. Patra, A. Amte, On the global trends and spread of the COVID-19 outbreak: preliminary assessment of the potential relation between location-specific temperature and UV index. J. Public Heal. pp. 1–10 (2020).

[29] J. Shaman, M. Kohn, Absolute humidity modulates influenza survival, transmission, and seasonality. Proceedings of the National Academy of Sciences 106, 3243–3248 (2009).

[30] E. Kudo, E. Song, L. J. Yockey, T. Rakib, P. W. Wong, R. J. Homer, A. Iwasaki, Low ambient humidity impairs barrier function and innate resistance against influenza infection. Proc. Natl. Acad. Sci. U. S. A. 166, 10905–10910 (2019).

[31] G. A. F. Seber, Multivariate observations, vol. 252 (John Wiley & Sons, 2009).

[32] B. Wang, L. Ho, Rainy season of the Asian-Pacific summer monsson. J. Clim. 15, 386–398 (2002).

[33] P. X. Wang, B. Wang, H. Cheng, J. Fasullo, Z. T. Guo, T. Kiefer, Z. Y. Liu, The global monsoon across time scales: Mechanisms and outstanding issues. Earth-Science Rev. 174, 84–121 (2017).

